# Undernutrition and associated factors among people living with HIV under NACS assessment in Muchinga Province, Zambia, 2019-2020

**DOI:** 10.1101/2022.03.31.22273149

**Authors:** Ebedy Sadoki, Constance Wose Kinge, Zikhona Jojozi, Grain Mwansa, Ben Chirwa, Frank Chirowa, Eula Mothibi, Thapelo Maotoe, George Magwende, Frank Shingwe, Ian Sanne, Philip Mwala, Charles Chasela

## Abstract

**Introduction:** Good nutrition in People Living with HIV (PLHIV) has a good influence on treatment outcomes and in turn a better quality of life. Despite, the significant role it plays, many patients have limited access to Nutritional Assessment Counselling and Support (NACS). We evaluated undernutrition in people living with HIV and associated factors in Muchinga Province, Zambia, from October 2019 to March 2020.

**Material and Methods:** This was secondary analysis of routine program data of HIV-positive clients on ART enrolled at EQUIP-supported health facilities in Muchinga. Undernutrition was determined using body mass index (BMI) calculations and classified as undernutrition (<18.5 kg/m^2^), normal (18.5 – 24.9 kg/m^2^) or over-nutrition (overweight, 25 – 29.9 kg/m^2^ and obese, 25 – 29.9 kg/m^2^). Multivariate-adjusted odds ratios (aOR) were used to assess factors associated with undernutrition.

**Results:** Of the 506 eligible clients under NACS, the mean age was 34.9 years ± 13.5SD, with 251 (approximately 50%) between the ages of 21 – 39 years. More than half (67%) were females, 284 (56%) were urban residents, and 180 (35.6 %) were unemployed. The majority (approximately 71%) were on the TLE regimen with a median duration on ART treatment of ∼3 years (IQR=1– 6). There were 233 (46%) who had a normal BMI, 191 (37.7%) who had under-nutrition, and 82 (16.2%) who had over-nutrition (9.7% overweight: 6.5% obesity). Clients in the urban area (aOR= 2.0; 95%CI: 1.28 – 3.1), unemployed (aOR= 2.4 (1.18-4.69)2.4; 95%CI: 1.18 – 4.69), married (aOR= 2.3; 95%CI: 1.26 – 4.38) and being on TLD (aOR= 2.8; 95%CI: 1.23 – 6.23) were more likely to be under-nourished.

**Conclusion:** NACS played a vital role in identifying HIV-positive clients who required more specialized care for improved clinical health outcomes. There is a need to strengthen HIV and nutrition integration in low-resourced countries with high HIV burden for improved treatment outcomes and quality of life.

## Introduction

Nutrition plays an important role in the management of people living with HIV (PLHIV). Good nutrition on PLHIV has a good influence on treatment outcomes and poses a better quality of life^1^. Human Immunodeficiency Virus (HIV) continues to be a significant global public health problem, with sub-Saharan Africa (SSA) being the most significantly affected^1, 2^. Sub-Saharan Africa contributes 70% (25.5 million) estimate of the global HIV infection burden and has a high burden of malnutrition^2, 3^. HIV care and antiretroviral therapy scale-up and coverage have played a significant role in AIDS-related deaths which have reduced by more than 51% since 2014^4^. Despite high ART scale-up and improved treatment outcomes, poor clinical outcomes remain high among PLHIV in both high and low resource settings due to compromised nutrition^5^.

The Government of Zambia began promoting interventions to improve the nutritional status of the PLHIV by making Nutrition Assessment and Counselling Services (NACS) mandatory to identify undernutrition status early, prevent undernutrition and improve treatment outcomes. EQUIP Zambia is implementing an integrated and HIV NACS program in three provinces (Muchinga, Luapula, and Northern). We evaluated undernutrition among PLHIV and determined its associated factors in Muchinga province.

## Material and Methods

### Study design and sites

This was secondary analysis of routine de-identified program HIV/NACS data in EQUIP-supported health facilities providing HIV care and treatment in Muchinga province, Zambia. Muchinga has a total of 148 facilities of which 48 are EQUIP-supported.

### Study population and data collection

The study population consisted of all people living with HIV and receiving ART care in all 48 EQUIP-supported facilities in Muchinga province. For logistic feasibility and inclusion criteria, only sites that contributed 60 percent of the total patients currently on treatment by March 31, 2020, were included in the study. All HIV positive clients who were clinically assessed for NACS and had complete anthropometric data were included in the analysis. De-identified clients’ demographics, clinical as well as related data collection included age, sex, residency, occupation, income, height, weight, duration on ART, and viral load. Weight was a measure to the nearest 1Kg, height was a measure to the nearest scale of 0.1cm using a (Salter scale). All information collected was recorded in the NACS log sheet.

### Data management and statistical analysis

Body mass index (BMI) was calculated as weight (kg)/height (m^2^) and categorized based on WHO guidelines as underweight (<18.5kg/m^2^), normal (18.5–24.9 kg/m^2^), overweight (25 – 29.9 kg/m^2^) and obese (25–29.9 kg/m^2^). For this study, undernutrition was defined as BMI below 18.5kg/m^2^. All analysis was performed using STATA/SE version 16.1 software (StataCorp, College Station, Texas 77845 USA). Categorical and continuous variables were summarized using proportions and median/means, respectively. Differences between proportions were determined using chi square test and Fischer exact test for sparce data. Multivariate logistics regression was used to identify factors associated with undernutrition.

### Ethical clearance

Since nutritional assessment forms a part of the HIV care and treatment services provided to PLHIV, no informed consent from clients was required. Permission to access de-identified client data was granted by Right to Care Zambia Not for Profit Organisation (NPO). Ethics was obtained from ERES CONVERGE IRB, Zambia (IRB No. 00005948).

## Results

### Characteristics of participants

Of the 7,898 clients assessed, only 506 (6.4%) were eligible for this study. The majority (n=7,392) were excluded due to inadequate documentation in the NACS logs sheet, no height or weight measurements and subsequent current nutrition status (Figure 1).

**Figure 1:**
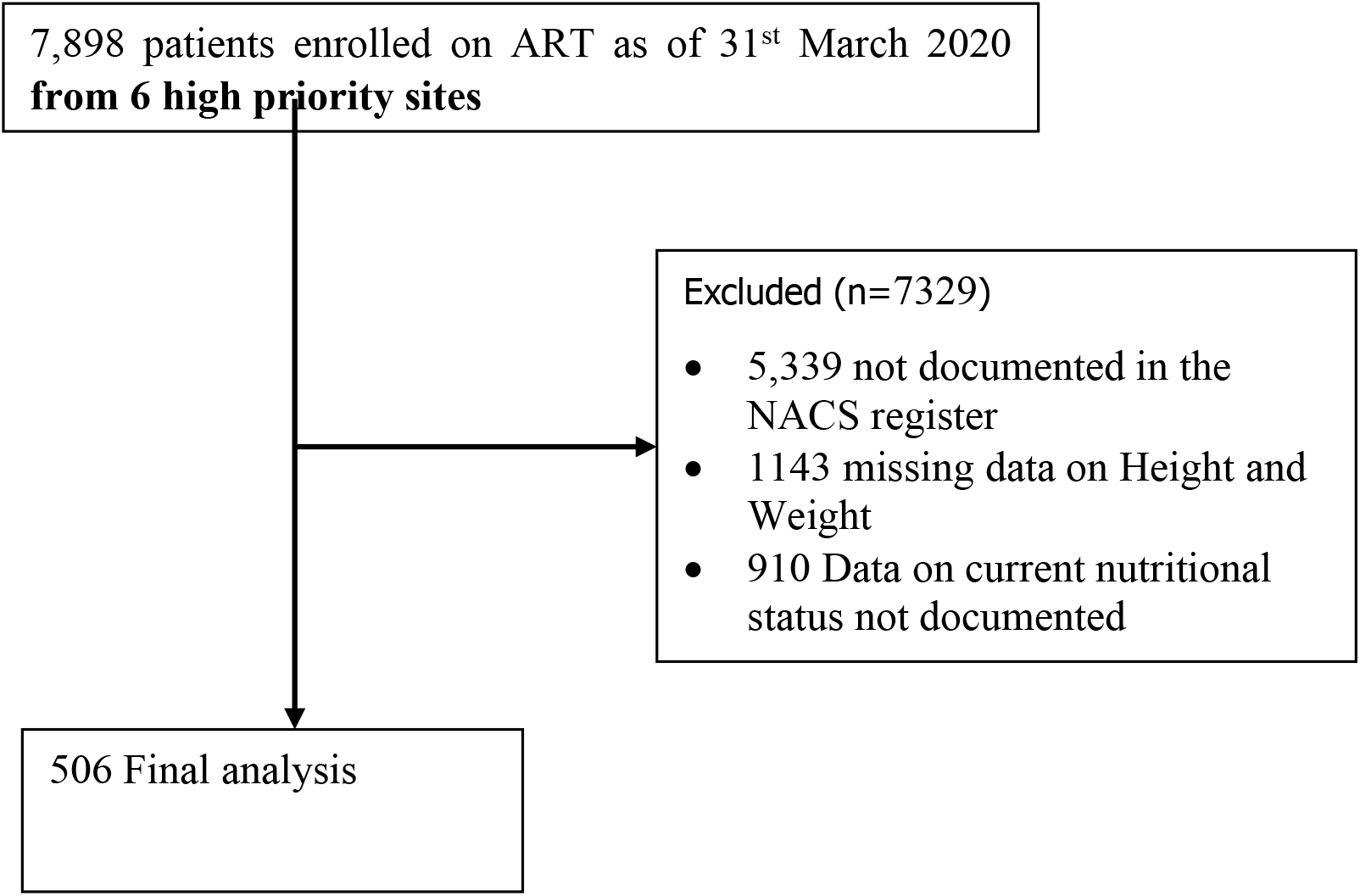
Participant Enrolment Flow

The mean age (SD) of the participants (n=506) was 34.9 (13.5) years. Most participants were female, unemployed, ever married, of low income, at least had a primary school education, suppressed viral load, on ART for less than 3 years, and were on TLE. Of the 506 participants, 191 (37.8%) were undernourished and the rest were either normal or overweight/obese (Table 1).

**Table 1.** Characteristics of the study population by Nutrition status

| Variables | Total (%) | Undernourished<br>N=191 (37.8%) | Normal<br>N=233 (46.1%) | Overweight<br>N=49 (9.7%) | Obese<br>N=33 (6.5%) |
| --- | --- | --- | --- | --- | --- |
| Age in years, mean (SD) | 34.9 (13.5) | 31.2 (14.9) | 36.0 (11.7) | 40.6 (11.0) | 40.5 (14.4) |
| Age group |  |  |  |  |  |
| ≤20 | 71 (14.0) | 51 (26.7) | 16 (6.9) | 2 (4.1) | 2 (6.1) |
| 21-39 | 251 (49.6) | 86 (45.0) | 135 (57.9) | 19 (38.8) | 11 (33.3) |
| 40 and above | 184 (36.6) | 54 (28.3) | 82 (35.2) | 28 (57.1) | 20 (60.6) |
| **Sex** |  |  |  |  |  |
| Male | 166 (32.8) | 63 (33.0) | 86 (36.9) | 10 (20.4) | 7 (21.2) |
| Female | 340 (67.2) | 128 (67.0) | 147 (63.1) | 39 (79.6) | 26 (78.8) |
| **Residency** |  |  |  |  |  |
| Rural | 222 (43.9) | 65 (34.0) | 108 (46.3) | 29 (59.2) | 20 (60.6) |
| Urban | 284 (56.1) | 126 (66.0) | 125 (53.7) | 20 (40.8) | 13 (39.4) |
| **Occupation** |  |  |  |  |  |
| Business | 77 (15.2) | 18 (9.4) | 42 (18.0) | 11 (22.5) | 6 (18.2) |
| Farmer | 117 (23.1) | 33 (17.3) | 62 (27.0) | 12 (24.5) | 10 (30.3) |
| Unemployed | 180 (35.6) | 98 (51.3) | 66 (28.3) | 7 (14.3) | 9 (27.3) |
| Unknown | 34 (6.7) | 7 (3.7) | 24 (10.0) | 3 (6.0) | 0 (0.0) |
| Other | 98 (19.4) | 35 (18.3) | 39 (16.7) | 16 (32.7) | 8 (24.2) |
| **Marital Status** |  |  |  |  |  |
| Single | 122 (24.1) | 54 (28.3) | 51 (21.9) | 9 (18.4) | 8 (24.2) |
| Married | 261 (51.6) | 109 (57.1) | 115 (49.4) | 22 (44.9) | 15 (45.5) |
| Divorced | 39 (7.7) | 10 (5.2) | 18 (7.7) | 8 (16.3) | 3 (9.1) |
| Widowed | 24 (4.7) | 4 (2.1) | 13 (5.6) | 4 (8.2) | 3 (9.1) |
| Unknown | 60 (11.9) | 14 (7.3) | 36 (15.4) | 6 (12.2) | 4 (12.1) |
| **Income** |  |  |  |  |  |
| <K500 | 26 (5.1) | 6 (3.1) | 18 (7.7) | 2 (4.1) | 0 (0.0) |
| K500 - K999 | 62 (12.3) | 17 (9.0) | 33 (14.2) | 10 (20.4) | 2 (6.1) |
| K1000 - K1499 | 23 (4.6) | 11 (5.8) | 6 (2.6) | 4 (8.2) | 2 (6.0) |
| K1500 - K1999 | 10 (2.0) | 1 (1.0) | 8 (3.4) | 1 (2.0) | 0 (0.0) |
| K2000 - K2999 | 11 (2.2) | 6 (3.1) | 4 (1.7) | 1 (2.0) | 0 (0.0) |
| >=K3000 | 36 (7.0) | 11 (6.0) | 16 (6.9) | 4 (8.2) | 5 (15.2) |
| Unknown | 338 (66.8) | 139 (73.0) | 148 (63.5) | 27 (55.1) | 24 (72.7) |
| **Educational Level** |  |  |  |  |  |
| Not yet in school | 15 (3.0) | 10 (5.2) | 3 (1.3) | 0 (0.0) | 2 (6.1) |
| Primary | 158 (31.2) | 78 (40.8) | 61 (26.2) | 11 (22.5) | 8 (24.2) |
| Secondary | 219 (43.3) | 62 (32.5) | 117 (50.2) | 26 (53.0) | 14 (42.4) |
| Tertiary | 29 (5.7) | 9 (4.7) | 12 (5.2) | 5 (10.2) | 3 (9.1) |
| Never been to school | 17 (3.4) | 8 (4.2) | 8 (3.4) | 0 (0.0) | 1 (3.0) |
| Unknown | 68 (13.4) | 24 (12.6) | 32 (13.7) | 7 (14.3) | 5 (15.2) |
| **Viral Load (copies/mL)** |  |  |  |  |  |
| <1000 | 275 (54.4) | 106 (55.5) | 131 (56.2) | 23 (47.0) | 15 (45.5) |
| ≥1000 | 35 (6.9) | 19 (10.0) | 11 (4.7) | 3 (6.1) | 2 (6.0) |
| Missing | 196 (38.7) | 66 (34.5) | 91 (39.1) | 23 (46.9) | 16 (48.5) |
| **ART Regimen** |  |  |  |  |  |
| TLD | 92 (18.2) | 42 (22.0) | 30 (12.9) | 12 (24.5) | 8 (24.0) |
| TLE | 357 (70.5) | 128 (67.0) | 175 (75.1) | 33 (67.3) | 21 (64.0) |
| Other | 57 (11.3) | 21 (11.0) | 28 (12.0) | 4 (8.2) | 4 (12.0) |
| **Years on ART** |  |  |  |  |  |
| < 3 | 254 (50.2) | 104 (54.4) | 115 (49.4) | 20 (40.8) | 15 (45.4) |
| 3 - <6 | 116 (22.9) | 44 (23.0) | 57 (24.5) | 10 (20.4) | 5 (15.2) |
| 6 - <9 | 82 (16.2) | 28 (14.7) | 36 (15.4) | 10 (20.4) | 8 (24.2) |
| 9 years and above | 54 (10.7) | 15 (7.9) | 25 (10.7) | 9 (18.4) | 5 (15.2) |

### Factors associated with undernutrition

In the bivariate analysis, age, residence, occupation, income, educational level and viral load were significantly associated with undernourishment. Age was protective with 80% less odds for undernutrition among age groups above 20 years. Similarly, marital status was protective against undernutrition for all categories of marriage except for those single. Area of residence (rural/urban), income, educational level and viral load were statistically significant with high odds for undernourishment. In the multivariable analysis, age was protective with 80% to 90% reduction in odds for undernutrition among those aged 21-39 years and above 40 years respectively. The odds of undernutrition were two times higher for urban residents, and among those unemployed. While marital status was protective in bivariate analysis, there was two times odds for undernutrition among those married compared to being single. While viral load was statistically significant in the bivariate analysis, there was borderline statistical significance with two times odds for undernutrition among those viral unsuppressed (Table 2).

**Table 2:**
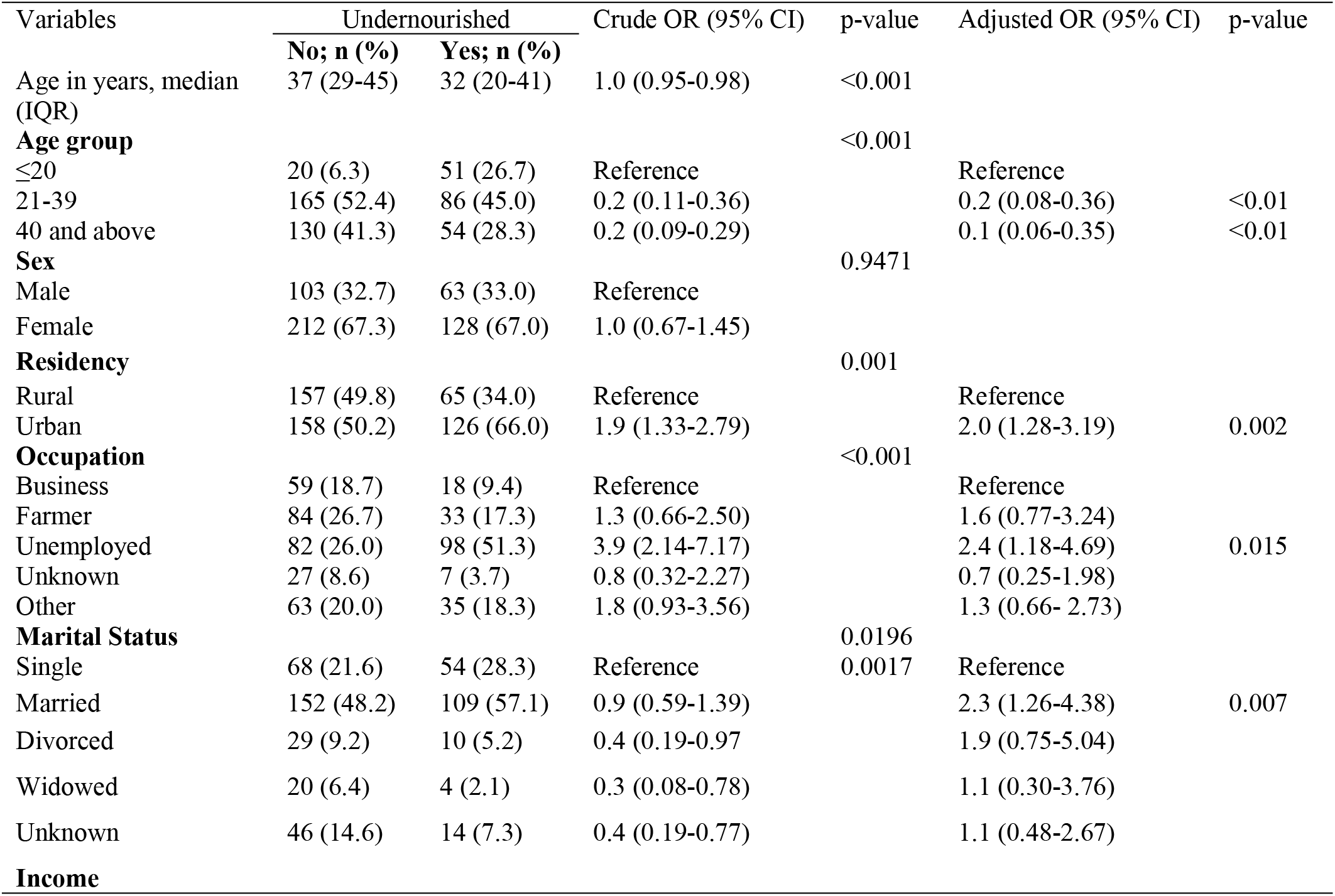
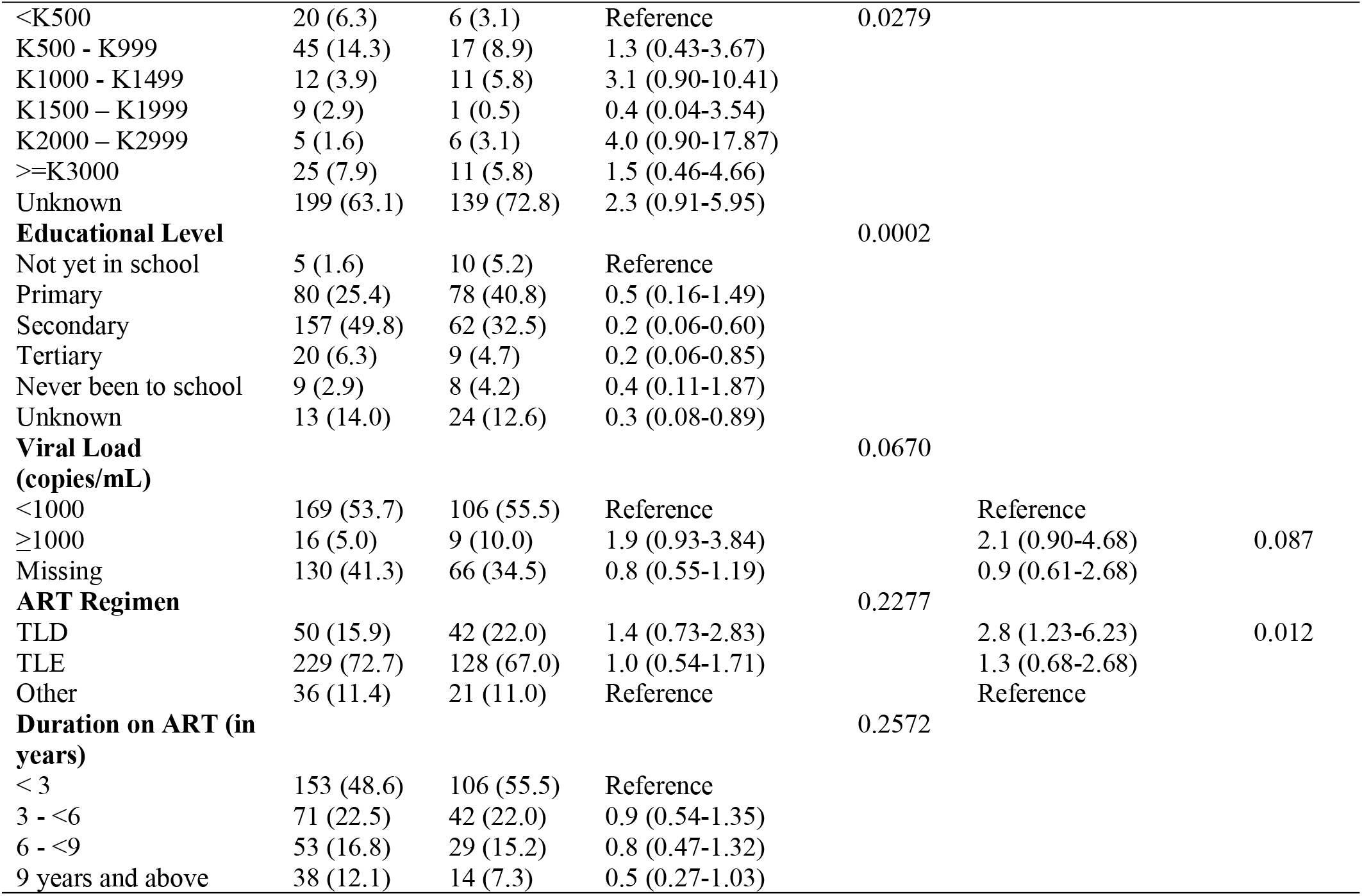
Factors associated with undernutrition

## Discussion

In this study, we evaluated undernutrition using NACS and determined its associated factors among PLHIV in Muchinga Province, Zambia. Undernutrition was found in more than one-third of the population with most either normal, overweight or obese. Most of the participants evaluated were middle aged, resident in urban, married, on TLE and less than 3 years on ART. Age was protective and being undernourished was less likely in the older age groups. However, being an urban resident, being married, unemployed, being on TLD and viral unsuppression increased the odds for undernutrition.

The protective age observed is in line with other studies that showed reduced chances of undernutrition among those who were older^6^. While you would expect undernutrition to be more in rural areas, our study found higher odds of undernutrition among those living in the urban areas. This may be attributed to food insecurity as opposed to those in the rural areas who engage in extensive farming activities and therefore have more access to food. In addition, the population evaluated were more from the urban population. In a qualitative study done in urban Zambia among women assessing acceptance and adherence to ART, hunger was found to be one of the factors affecting adherence and ART intake. In another food insecurity report, 93% of households in the informal settlements, (making 75% of Lusaka’s urban population) were food insecure^7^ affirming nutrition vulnerabilities in some urban sub-populations. Food insecurity has been found to be a major cause of undernutrition, thus compromising treatment outcomes among PLHIV^8, 9^.

In our study, those on TLD regimen had three times odds of being undernourished than those on other regimens. This is different with other studies where the transition to DTG in virally suppressed adolescents (aged 10–19 years) was associated with an increase in the rate of BMI change^10^. In this study, though unsuppressed viral load was shown to be borderline statistically significant with two times odds, this finding is consistent with a study in Malawi that showed unsuppressed viral load and undernutrition among many of the factors associated with high mortality among PLHIV on ART^11^.

In conclusion Nutritional Assessment, Counselling, and Support played a vital role in identifying HIV-positive clients who require more specialized care for improved clinical health outcomes. This is more crucial especially during this era of COVID-19. There is therefore a need for continued strengthening of HIV and nutrition integration in countries with a high HIV burden to prevent treatment disruption for improved quality of life. NACS helps with the prevention of undernutrition and early identification and treatment, which leads to improvement of health outcomes for PLHIV in care.

## Data Availability

All relevant data are within the manuscript and its Supporting Information files

## Acknowledgements

The authors wish to express sincere gratitude to colleagues from the Ministry of Health Zambia and Right to Care Zambia Direct Service Delivery (DSD) staff for their technical support.

## Data Availability Statement

All relevant data are within the manuscript and its Supporting Information files.

